# Founder Mutation Effect Seen By *CERKL* Gene Mutation Causing Retinal Dystrophy in North Indian Population

**DOI:** 10.1101/2023.11.09.23298139

**Authors:** Mayank Bansal, Debojyoti Chakraborty

**Affiliations:** CSIR - Institute of Genomics and Integrative Biology, Delhi; Sightgenics Research, Delhi; Fortis Memorial Research Institute, India

**Keywords:** Founder mutation, *CERKL* gene mutation, genotype phenotype correlation, Inherited Retinal Dystrophy (IRD), Retinitis Pigmentosa (RP)

## Abstract

**Purpose:** This paper describes the clinical features, genotype phenotype correlation of *CERKL* gene mutation, one of the most common genetic mutations of Inherited Retinal Dystrophy (IRD) patients seen in our cohort in North India.

**Materials and Methods:** Patients clinically diagnosed with an IRD were included in the study. Patients underwent ultra widefield (UWF) fundus photographs, fundus autofluorescence (FAF), optical coherence tomography (OCT). A pedigree charting was done. Genetic testing by next generation sequencing (NGS) was done, clinical exome was analyzed.

**Results:** We report ophthalmic and genetic findings of seven patients with *CERKL* gene mutation of the thirty five patients that chose to undergo genetic sequencing (amongst our cohort of sixty two patients with IRD). The age ranged from 17 to 45 (median 25) years. Vision ranged from LogMAR 0.18 to 1.8. OCT showed central macular thickness (CMT) ranging from 103 to 268 microns. Majority patients’ fundus exhibited macular pigmentary changes with atrophy, paucipigmentary or limited peripheral retinal pigmentary changes; mild optic disc pallor, and minimal vascular attenuation. Stippled hypo autofluorescence at the macula was the most common finding, with minimal hypoautoflorescence in the retinal periphery.. The genetic sequencing of all patients showed the same mutation, a 2 base pair deletion in exon 7 of the *CERKL* gene (chr2:g.181548785_181548786del). Coincidentally, all patients with *CERKL* gene mutation were found to be from a single ethnic community suggesting a founder mutation effect.

**Conclusions:** Mutation in the *CERKL* gene results are among the most common causes of IRD in North India. Affected patients showed a definitive early macular involvement. This study reports the presence of a founder mutation effect in the *CERKL* gene in a large ethnic community in North India.

## Introduction

While globally, the prevalence of Inherited Retinal Dystrophies (IRDs) including Retinitis Pigmentosa (RP) is around 1 in 4000, the rate is significantly higher in India. (1) One comprehensive survey in India showed that in rural areas, 1 in 372 people is affected by RP, while in urban regions the ratio is 1 in 930. (2) Additionally, a population-oriented, cross-sectional study conducted in Central India evaluated the fundus photos of 4,543 individuals and found that 1 in 750 individuals in rural Central India are affected by RP. (3) Extrapolating from this data, it’s estimated that half a million people in India have RP, and about 1.4 million carry the genetic mutations linked to the disease. Another study from South India, which conducted eye examinations on 2,513 patients visiting a specialized eye hospital, found that out of 673 patients with genetic eye disorders, 430 had RP. These RP cases predominantly showed high levels of consanguinity and followed an autosomal recessive inheritance pattern. (4)

The use of Next-Generation Sequencing (NGS) to identify causative mutations in IRD patients has seen a steady rise. This method has been instrumental in understanding the unique distribution of causative mutations, worldwide, also helping understand geographic distribution of IRD mutations. In our practice we see patients primarily from North India where we found mutations in the *CERKL* gene to be among the more common mutations which we sought to describe in this paper.

The *CERKL* gene produces ceramide kinase-like protein, a homologue of ceramide kinase (CERK) which is involved in the transformation of ceramide into ceramide-1-phosphate. It exists in seven alternatively spliced isoforms. The CERKL protein comprises of 558 amino acids and is predominantly found in the cytoplasm and nucleus, with notable enrichment in nucleoli. Ceramide itself is a metabolic byproduct of sphingophospholipids, an essential component of cell membranes. It plays a role in internal cell signaling and programmed cell death, or apoptosis. In simpler organisms, *CERKL* expression has been observed mostly in the Ganglion Cell Layer (GCL), and to a lesser degree in the Outer and Inner Nuclear Layers (ONL and INL, respectively). (5)

There is growing evidence suggesting that CERKL, through its involvement in sphingophospholipid and ceramide metabolism, serves to protect photoreceptor cells from oxidative stress. When CERKL is not functioning correctly, photoreceptor cells become susceptible to apoptosis, resulting in significant degradation of both rod and cone cells. (6) Tuson and colleagues discovered a specific homozygous nonsense mutation, p.Arg257ter (c.769 C>T), in exon 5 of the *CERKL* gene, to be cause of autosomal recessive RP26. (7) Sengillo et al described four patients with *CERKL* gene mutation. In their study, reduced visual acuity and early onset of maculopathy was seen in all individuals. They further described peripheral retinal punched out degenerative lesions, and numerous hyperautofluorescent foci encircling a region of central atrophy at the macula. Patients had mutations in the following - Homozygous c.847□JC□J>□JT (p.R283*) in two patients, c.769□JC□J>□JT (p.R257*) in one patient, and c.1303□JC□J>□JT (p.R435*) in one patient. (8) Sen et al described a novel variant, c.899-IG>A in the *CERKL* gene causing IRD. (9) Most recently, Varela et al described *CERKL* variants from the United Kingdom. THey found that the *CERKL*-retinal dystrophy phenotype varies widely, spanning from isolated macular conditions to extensive peripheral retinal involvement, accompanied by diverse functional traits that don’t completely align with the typical rod-cone or cone-rod classifications. (10)

The above studies give an excellent description of *CERKL* gene mutation causing IRDs. Further, we found that mutations in the *CERKL* gene were among the most common mutations in our cohort of sequenced IRD patients. To add, all patients in our series, while not consanguineous, belonged to the same ethnic community. Therefore, in this descriptive study, we aim to describe the phenotypic characteristics of these patients.

## Methods

Patients visiting our practice as well as seeking teleconsultation between 2019 to 2023 with findings indicating IRD, were given the option for genetic testing and guidance. Patients were typically advised clinical exome sequencing, using next generation sequencing (NGS) protocol. Of the thirty five patients that chose to undergo genetic sequencing, seven were found to have *CERKL* gene mutation, and were included in this study.

### Clinical Methods

A cross-sectional analysis of 14 eyes of these seven unrelated patients who were diagnosed with *CERKL* gene mutation was done. We reviewed patient phenotype information which included both clinical data and any supplementary tests carried out during visits to our practice. Demographics including ethnicity of patients was recorded. Detailed history including pedigree charting was reviewed. Ethics approval was obtained from the institutional review board and study done according to the tenets of declaration of Helsinki. Factors such as the clinical symptoms, age at which symptoms began, and any history of difficulty seeing in low light were documented. Metrics like Best Corrected Visual Acuity (BCVA) were measured using Snellen’s charts for both distance and near vision. The refraction was recorded as a spherical equivalent, and findings from slit-lamp biomicroscopy and indirect ophthalmoscopy were also included.

Images of the retina were captured using either the Carl Zeiss FF450 IR camera or ultra-widefield pseudocolor imaging via Optos device. These images helped in documenting various phenotype characteristics, such as optic disc pallor, thinning of arteries, irregularities in the Retinal Pigment Epithelium (RPE), and any macular changes. The Optos device was also used for Fundus Autofluorescence (FAF). Findings from Optical Coherence Tomography (OCT), using either Carl Ziess Cirrus OCT, or Optovue RTVue OCT, were included in the records. Visual Fields records were analyzed when available, which when done were on the Humphrey or Octopus Visual Field Analyzer platforms. Electroretinogram (ERG) records were also reviewed when available.

### Genetic Screening (Next Generation Sequencing)

For patients a comprehensive family history was charted. After taking informed consent from the patient or family members, a peripheral blood sample (3 ml) was collected into an EDTA vacutainer. Genomic DNA was then isolated, which underwent targeted gene capture through a specialized capture kit. These genetic libraries were then sequenced to an average depth of 80-100X using the Illumina sequencing platform. The generated sequences were aligned to the human reference genome (GRCh38) using the BWA aligner, part of the Sentieon suite. Sentieon was further employed for tasks like duplicate removal, indel recalibration, and realignment.

The GATK best practices framework for the identification of germline variants in the sample using Sentieon was followed. The sequences that were obtained were aligned to the human reference genome (GRCh38) using the BWA aligner and were analyzed using Sentieon for the removal of duplicates, recalibration, and re-alignment of indels. Sentieon haplotype caller was then used to identify variants in the sample. The germline variants that were identified in the sample were deeply annotated using the VariMAT pipeline. Gene annotation of the variants was performed using the VEP program against the Ensembl release 99 human gene model. In addition to SNVs and small indels, copy number variants (CNVs) were detected from targeted sequence data using the ExomeDepth method. This algorithm detected CNVs based on a comparison of the read-depths in the sample of interest with the matched aggregate reference dataset.

Clinically relevant mutations in both coding and non-coding regions were annotated using published variants from scientific literature and a selection of disease-related databases, such as ClinVar, OMIM, HGMD, LOVD, DECIPHER (for population CNVs), and SwissVar. Common variants were screened out based on their allele frequency using databases like 1000Genome Phase 3, gnomAD (versions 3.1 & 2.1.1), dbSNP, 1000 Japanese Genome, TOPMed (Freeze_8), Genome Asia, and the in-house Indian population database, MedVarDb v2.1.

The impact of non-synonymous variants was assessed through various computational algorithms, including PolyPhen-2, SIFT, MutationTaster2, and LRT. Variants determined to be of clinical significance were then chosen for further interpretation and included in the final reporting.

## Results

Mutations in *CERKL* gene were identified in seven patients, whose age ranged from 17 to 45 years at first diagnosis (Table 1). There were two female and five male patients. There was no history of consanguinity in any of the patients, as well as no family history of progressive non treatable vision loss. All the same, all patients with the *CERKL* gene mutation had ancestry in North India and belonged to the same ethnic group, of the Agarwal community.

Best Corrected Visual Acuity (BCVA) of the patients ranged from LogMAR 0.18 to 1.8. It’s worth mentioning here that the majority of patients (6 of 7) had vision ranging from LogMAR 0.18 to 0.50 in the better eye. One patient however, deteriorated from LogMAR 0.5 (20/60) to 1.7 (counting finger close to face) vision over a period of 3 years, between the age of 17 to 19 years. The anterior segment examination was unremarkable for all patients, clear cornea and lens.

Fundus examination showed the following findings. The macula for 12 eyes of 6 patients showed stippling and atrophic changes. Both eyes of a single patient showed minimal thickening. None of the patients had epiretinal membrane formation, or frank cystoid macular edema (CME). There were no full thickness, lamellar or pseudo macular holes seen.

The optic disc in 3 of 7 patients had normal healthy appearance, with disc cupping ranging from 0.3 to 0.5:1 and pink healthy neuroretinal rim (NRR), distinct clear disc margins. Rest of patients (4 of 7), had minimal disc pallor, normal cupping, with the NRR being mildly pale as compared to a normal healthy looking disc. None of the patients had classic waxy disc pallor, optic atrophy appearance, or gliosis over the optic disc.

The peripheral retina in the majority of patients (6 of 7) showed minimal pigmentary changes. There were paucipigmentary changes, with a thinned out appearance of retina outside the arcades as compared to the posterior pole, but there were no no bony spicules. Only one patient (1 of 7) had a classic retinitis pigmentosa (RP) pattern of bony spicules.

The retinal vasculature in 2 of 7 patients did not show any changes, vessels had normal caliber, with no attenuation. Rest of patients (5 of 7) showed minimal arteriolar attenuation. None of the eyes had significant arteriolar attenuation classically described in RP.

The Fundus Autofluorescence (FAF) was instrumental in revealing underlying changes, especially when there were negligible pigmentary changes in retinal periphery. The patients with pauci-pigmentary changes, revealed hypo autofluorescence in the retinal periphery, while the central macula showed stippled autofluorescence. In the patient with obvious bony spicules the FAF showed marked hypo autofluorescence in the retinal periphery and stippled autofluorescence at the macula. The degree of stippling at the macula was commensurate with the amount of vision loss.

The OCT in the majority of patients (6 of 7) showed thinning at the fovea, with loss of outer retinal layers, including the ellipsoid zone. The central macular thickness ranged from 123 to 268 microns, mean of 210.71 microns. One patient had minimal foveal thickening, with loss of foveal dip, with CMT of 243 microns. None of the patients had epiretinal membrane formation, or frank cystoid macular edema (CME). There were no full thickness, lamellar or pseudo macular holes seen, commensurate with clinical findings.

Of special mention here is a male patient, age ranging between 16-20 years; with significant vision loss (OD hand motion vision and OS counting finger vision). This patient had minimal peripheral pigmentary changes, clinical periphery had an appearance of early fundus albipunctatus. While the macula had significant atrophic pigmentary changes. The FAF showed marked areas of hypo autofluorescence in macula, while the periphery had minimal hyper autofluorescence areas. The OCT had corresponding marked foveal atrophy with CMT of 123 and 133 microns. This patient had shown rapid deterioration of vision loss over three years from 6/18 in the better eye to counting fingers. This one patient did not have any other underlying ocular or systemic co-morbidity, there were no environmental or personal history factors (smoking, alcohol, drug abuse) which could explain the rapid visual loss.

Electroretinogram (ERG) was done for two patients, where it demonstrated a reduced photopic and scotopic response bilaterally, despite minimal pigmentary changes, suggesting both rods and cones being affected. The vision in both these patients ranged from 20/40 to 20/60. Visual Fields (Humphrey Visual Field Analyser), 30-2 SITA standard protocol, was available for two patients, both of whom had vision between 20/40 to 20/60. The fields showed peripheral field loss, with reduced central sensitivity.

The genetic sequencing of all patients showed the same mutation, a 2 base pair deletion in exon 7 of the *CERKL* gene (chr2:g.181548785_181548786del) that results in a frameshift and premature truncation of the protein 20 amino acids downstream to codon 323 (p.Met349ValfsTer20; NM_201548.5) (The observed variant has previously been reported as c.1045_1046del based on the transcript NM_001030311.2). All patients were homozygous for this mutation, with autosomal recessive inheritance. None of the patients had more than one IRD causing gene mutation. All patients were non-syndromic.

To look for any subtle retinal changes in carrier progeny of the patients, we reviewed records of the daughter of one of the affected patients, who was a carrier for the same *CERKL* gene mutation. Her vision was 20/20 both eyes, with no changes in the fundus, and the FAF was unremarkable (figure).

## Discussion

More than 20 distinct *CERKL* transcripts have been identified in the human retina, primarily due to extensive alternative splicing and numerous translational start sites. (11) Interestingly, findings from a double-knockout zebrafish model indicated that rods experienced earlier and more pronounced degeneration compared to cones.(12) However, in a knockdown-knockout mouse model, cones were found to be more severely affected.(13) Both models, however, concurred on the presence of abnormalities in photoreceptor outer segments, characterized by accumulation in the interphotoreceptor matrix and the elongated and disordered appearance of photoreceptor outer segments, which also perhaps suggests reduced phagocytic activity by the retinal pigment epithelium(RPE). Recent research has also shown that *CERKL* gene may provide protection against oxidative stress within RPE cells, suggesting that diminished RPE function may occur in its absence.(14)

The purpose of this study was to report the clinical findings in the most common mutation seen in our cohort of patients with IRD, i.e. mutations in the *CERKL* gene. Patients typically presented in the second decade, with two line vision loss. Further, we found all patients affected by this *CERKL* gene mutation to belong from the same large ethnic group, primarily originating in north India, and belonging to the Agarwal community, therefore having a founder mutation effect.

Founder mutations are genetic alterations that originate from a single common ancestor and are subsequently passed down to a significant portion of a population. These mutations have a pronounced effect on the genetic makeup of specific communities or populations. The described mutation in *CERKL* gene, was seen in the Agarwal community, with no history of consanguinity. This large ethnic group has ancestry primarily in North India. Incidentally, such founder effect in IRD due to *CERKL* gene mutations has also been reported in Finnish and Yemenite Jew populations. (15), (16)

Founder mutations can provide valuable insights into the study of genetic diseases, as they offer a clear genetic marker for researchers to investigate and develop targeted diagnostic and therapeutic strategies. Understanding the effects of founder mutations is essential for both medical genetics and population genetics, as it helps unravel the intricate relationship between genetics, ancestry, and inherited diseases in specific groups of people.(17)

Given the variable phenotype and expressivity of the same mutation, it’s plausible that modifier genes and complex genetic and environmental interactions may be at play. Additionally, certain other factors may interfere with *CERKL* gene’s interactions with other proteins, disrupting its intricate functional and regulatory roles, potentially affecting rods and cones differently. (10) Further investigations in the form of functional studies hold the potential to provide deeper insights into this subject.

It is crucial to establish these genotype-phenotype associations in IRDs. These associations are particularly possible to identify for retinal dystrophies, due to our ability to examine and document changes in the retina directly. The phenotype serves as a genetic signature imprinted on the retina. Furthermore, genetic testing, genomic sequencing for patients with IRDs has threefold benefits. Correlating the phenotype of IRD with the causative mutations, helps understand likely progression of the retinopathy, and therefore discuss prognosis with the affected family. Further, where relevant, it helps counsel the family regarding family planning, and chances of transmission in future generations. But most importantly, with the changing landscape of gene augmentation and editing therapies, it helps make patients aware which potential clinical trial they may be eligible for.(18) Our paper highlights the common genetic mutation from the Indian subcontinent, which lays ground for development of future therapies.

## Conclusion

This paper describes the *CERKL* gene mutation, having a founder mutation effect for IRDs in the Agarwal ethnic group, originating from North India. The said mutation leads to vision loss typically presenting in the second decade with early macular involvement, stippled pigmentary changes at macula, and limited pauci-pigmentary changes in retinal periphery.

## Data Availability

All data produced in the present study are available upon reasonable request to the authors

---

**Figure1:**
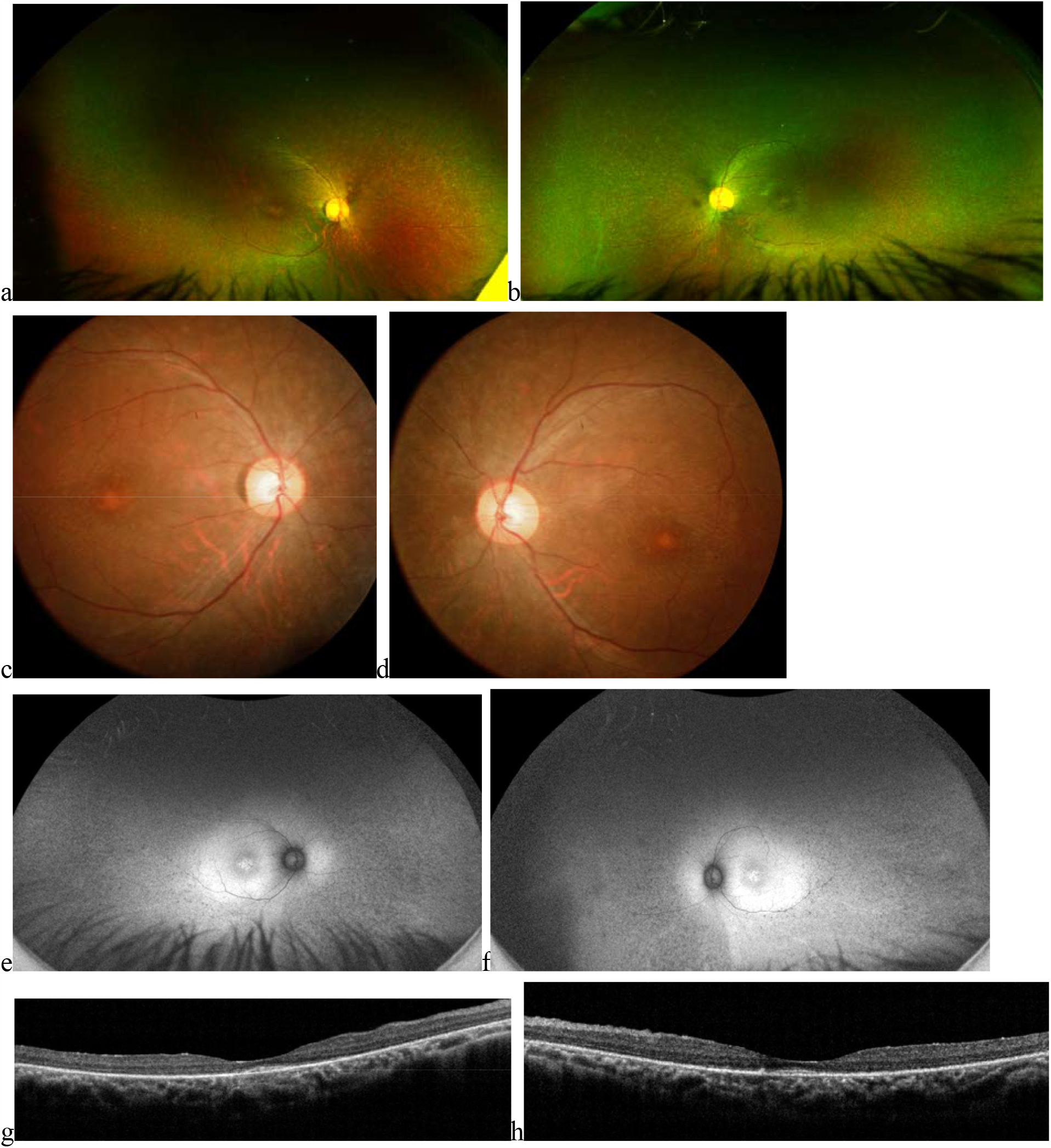
Ultrawide field (UWF) retinal image (a & b), fundus photo (c & d), fundus autofluorescence (FAF) image (e & f), and Optical Coherence Tomography (OCT) (g&h) of a male patient, aged 15-20 years, with Inherited Retinal Dystrophy (IRD), affecting by mutation in the *CERKL* gene. Note the paucipigmentary appearance in retinal periphery and atrophic changes at the macula. The FAF shows central stippled, hypo-autofluorescence and periphery hypo autofluorescence. Patient had a vision of 6/9p (LogMAR 0.18) in the right eye and 6/9 (LogMAR 0.18) in the left eye. Central macular thickness (CMT) measured 216 and 137 micrometers in right and left eye respectively. Notice the loss of outer retinal layers, ellipsoid zone on OCT

**Figure2:**
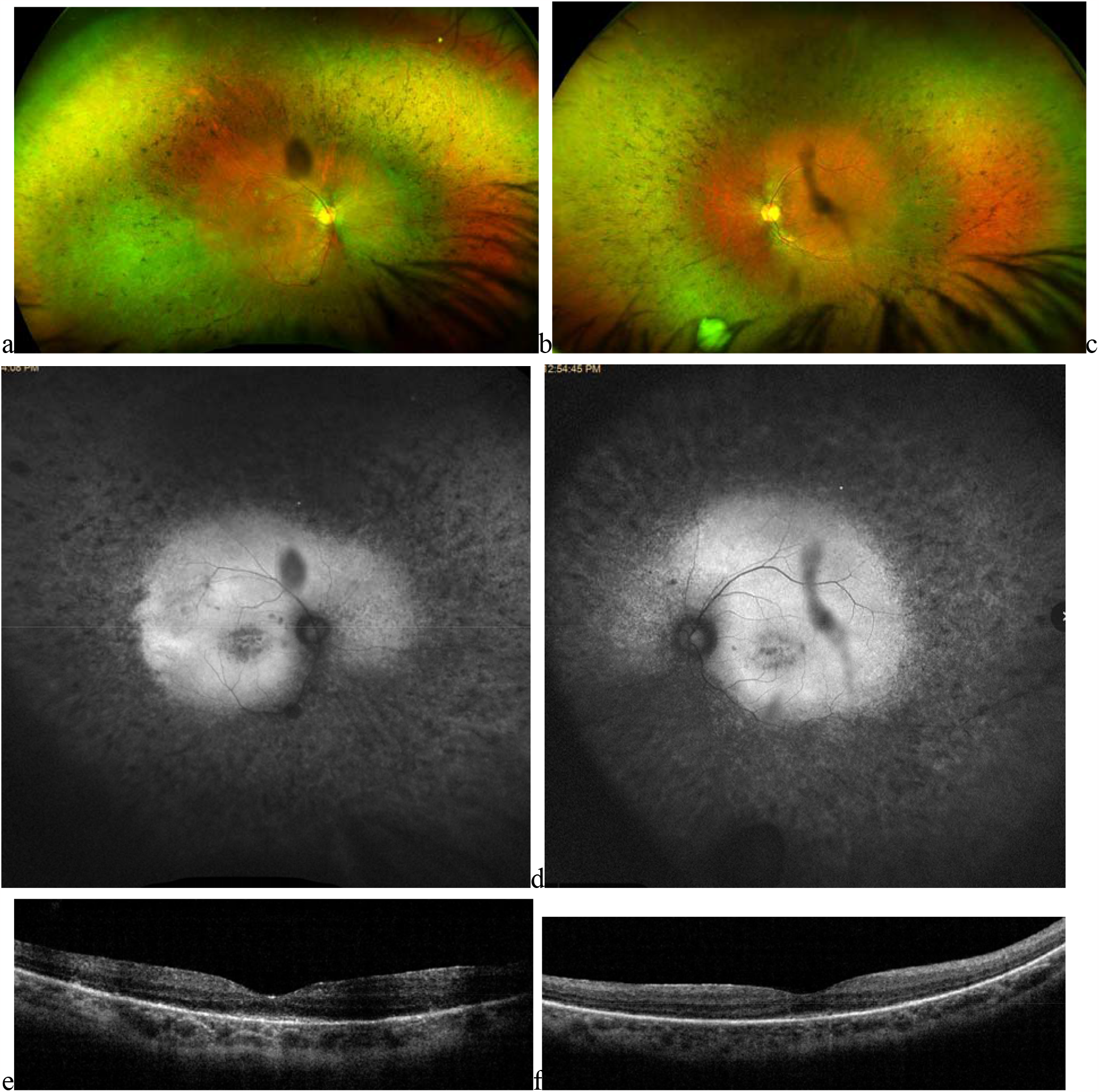
Ultrawide field (UWF) retinal image (a & b), fundus autofluorescence (FAF) image (c & d), and Optical Coherence Tomography (OCT) (e&f) of a male patient aged between 31-35 years with Inherited Retinal Dystrophy (IRD), affecting by mutation in the *CERKL* gene. The pigmentation with bony spicules in retinal periphery are classic of RP. The FAF shows central stippled autofluorescence and periphery hypo autofluorescence. Patient had a vision of 6/9p (LogMAR 0.18) in the right eye and 6/9 (LogMAR 0.18) in the left eye. Central macular thickness (CMT) measured 268 and 258 microns in right and left eye respectively. Notice the loss of outer retinal layers, ellipsoid zone on OCT at fovea.

**Figure3:**
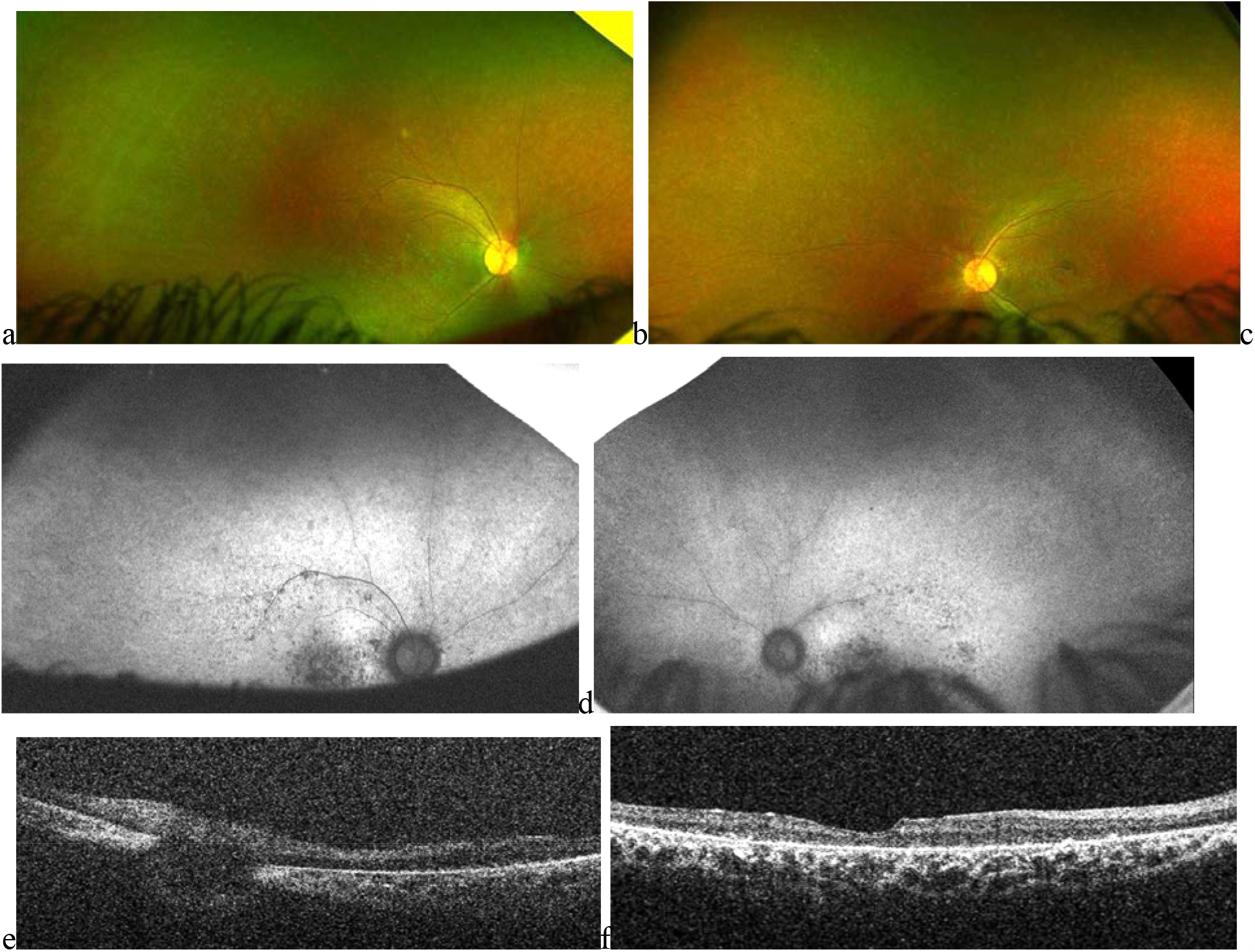
Ultrawide field (UWF) retinal image (a & b), fundus autofluorescence (FAF) image (c & d), and Optical Coherence Tomography (OCT) (e&f) of a male patient aged between 16 to 20 years with mutation in the *CERKL* gene. Patient had a vision of hand motions in the right eye and finger counting in the left eye. The retinal periphery has a paucipigmentary appearance. The FAF shows central hypo-autofluorescence and peripheral autofluorescence shows minimal changes, signifying primarily cone damage. Central macular thickness (CMT) measured 123 and 133 micrometers in right and left eye respectively, with a prominent loss of outer retinal layers.

**Figure4:**
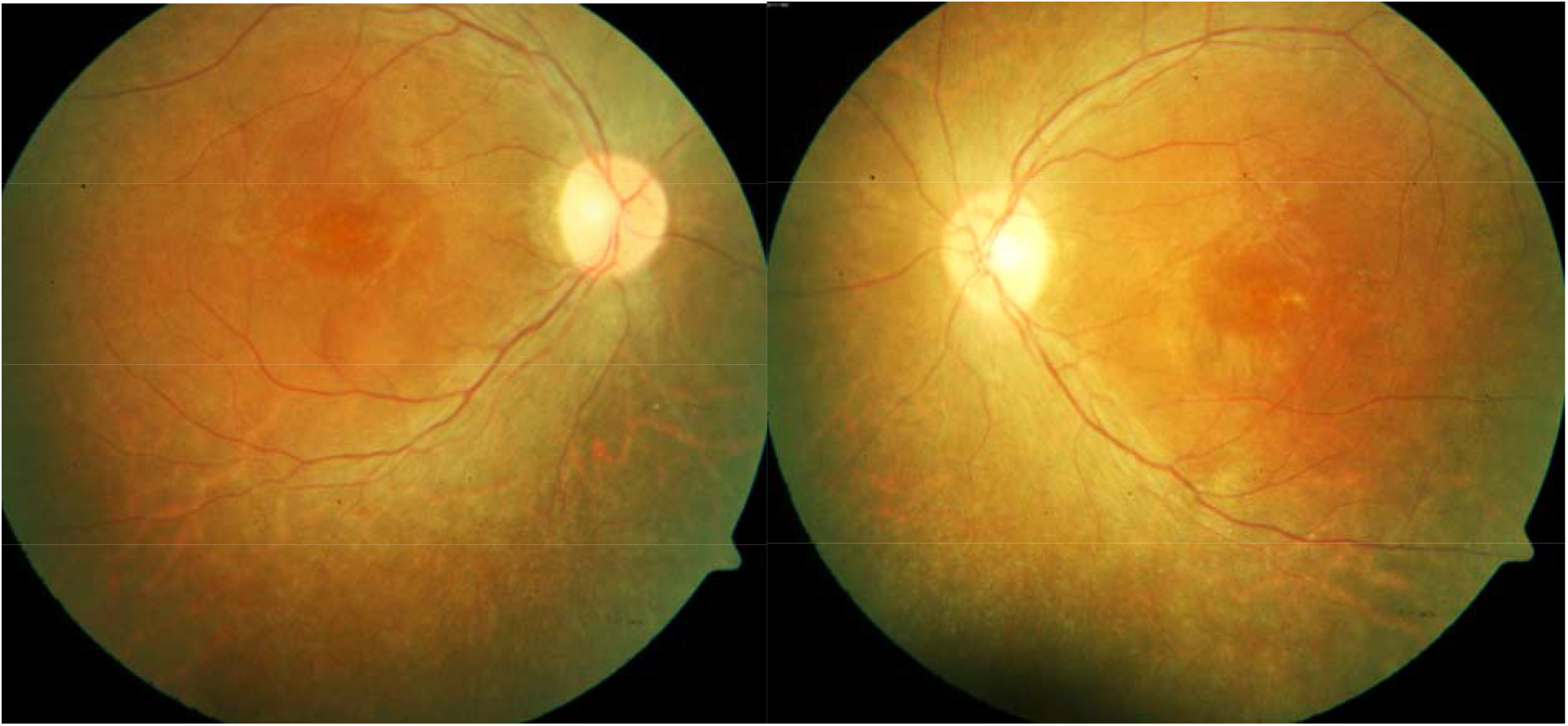
Fundus photo of a female patient, 21-25 years old, with mutation in the *CERKL* gene. Patient had a vision of 6/12 and 6/9 (LogMAR 0.30 and 0.18) respectively in the right and left eye. The pigmentation in the retinal periphery was sparse with a paucipigmentary appearance, and macula showed atrophic changes confirmed on OCT.

**Figure5:**
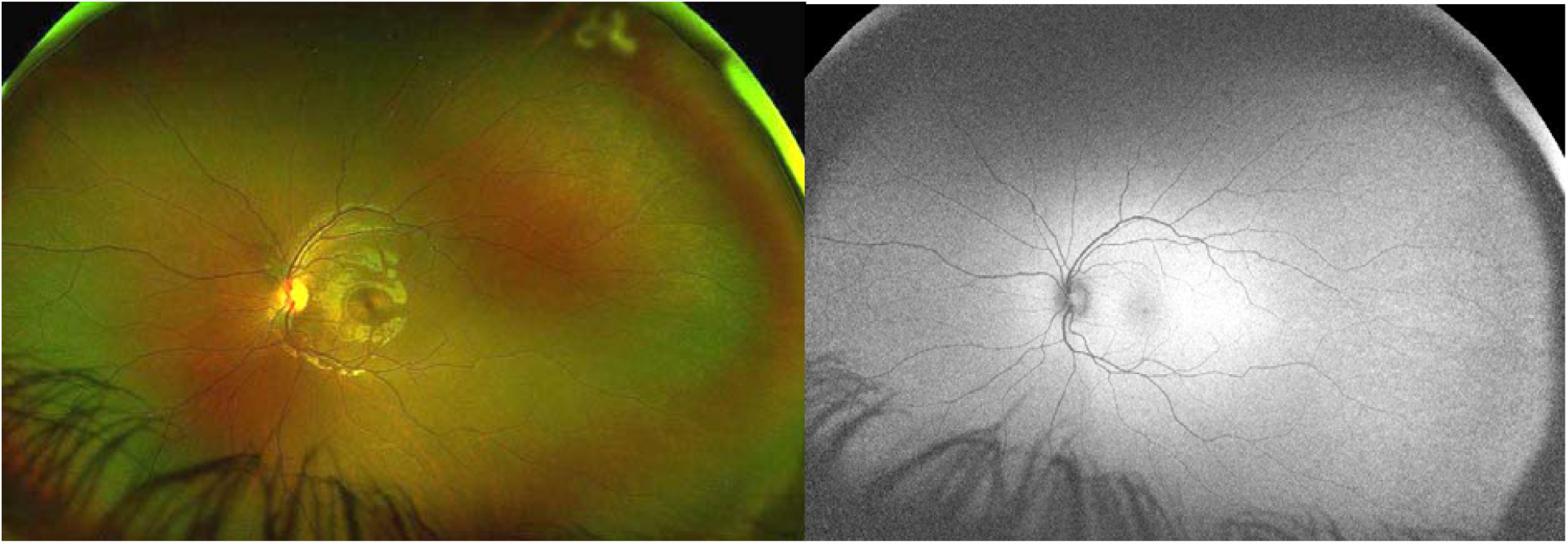
Ultra Wide field fundus photo and autofluorescence image of a carrier for the IRD causing *CERKL* gene mutation; female, 16-20 years; Both photographs appeared essentially normal, the patient had 20/20 vision and there were no changes on OCT.

