## Supplemental Table 1 for "Founder Mutation Effect Seen By *CERKL* Gene Mutation Causing Retinal Dystrophy in North Indian Population"

Table 1. Demographics and clinical findings of patients with IRD due to *CERKL* gene mutation

| *Age Range* | *Sex* | *LogMAR* | | *macula* | *Periphery* | *Optic disc* | *Fundus vessels* | *FAF* | *OCT OD* | *OCT feature OD* | *OCT OS* | *OCT feature OS* |
| --- | --- | --- | --- | --- | --- | --- | --- | --- | --- | --- | --- | --- |
| 21-25 | F | 0.30 | 0.18 | atrophic changes | pauci-pigmentary changes | mild disc pallor | minimal vascular attenuation |  | 221 | outer retinal loss | 200 | outer retinal loss |
| 41-45 | M | 0.30 | 1.00 | atrophic changes | pauci-pigmentary changes | mild disc pallor | minimal vascular attenuation |  | 209 | outer retinal loss | 216 | outer retinal loss |
| 31-35 | F | 0.30 | 0.30 | atrophic changes | pauci-pigmentary changes | normal disc appearance | minimal vascular attenuation |  | 257 | outer retinal loss | 256 | outer retinal loss |
| 21-25 | M | 0.5 | 1.5 | minimal thickening | pauci-pigmentary changes | normal disc appearance | no change |  | 243 | minimal thickening, single small cyst | 213 | minimal thickening |
| 16-20 | M | 0.18 | 0.18 | atrophic changes | pauci-pigmentary changes | normal disc appearance | minimal vascular attenuation | macular stippled autoflourescence; peripheral hypoautoflourescence | 216 | outer retinal loss | 137 | outer retinal loss |
| 36-40 | M | 0.18 | 0.18 | atrophic changes | bony spicules | mild disc pallor | minimal vascular attenuation | macular stippled autoflourescence; peripheral hypoautoflourescence in areas of bony spicules | 268 | outer retinal loss | 258 | outer retinal loss |
| 16-20 | M | 1.8 | 1.7 | atrophic changes | pauci-pigmentary changes | mild disc pallor | minimal vascular attenuation | macular stippled autoflourescence, peripheral minimal changes | 123 | outer retinal loss | 133 | outer retinal loss |
